# Outbreak dynamics of COVID-19 in China and the United States

**DOI:** 10.1101/2020.04.06.20055863

**Authors:** Mathias Peirlinck, Kevin Linka, Francisco Sahli Costabal, Ellen Kuhl

**Affiliations:** Departments of Mechanical Engineering and Bioengineering, Stanford University, Stanford, California, USA; Department of Mechanical and Metallurgical Engineering, School of Engineering · Institute for Biological and Medical Engineering, Schools of Engineering, Medicine and Biological Sciences, Pontificia Universidad Catolica de Chile, Santiago, Chile

**Keywords:** coronavirus, COVID-19, epidemiology modeling, SEIR model, network model

## Abstract

On March 11, 2020, the World Health Organization declared the coronavirus disease 2019, COVID19, a global pandemic. In an unprecedented collective effort, massive amounts of data are now being collected worldwide to estimate the immediate and long-term impact of this pandemic on the health system and the global economy. However, the precise timeline of the disease, its transmissibility, and the effect of mitigation strategies remain incompletely understood. Here we integrate a global network model with a local epidemic SEIR model to quantify the outbreak dynamics of COVID-19 in China and the United States. For the outbreak in China, in n = 30 provinces, we found a latent period of 2.56±0.72 days, a contact period of 1.47±0.32 days, and an infectious period of 17.82±2.95 days. We postulate that the latent and infectious periods are disease-specific, whereas the contact period is behavior-specific and can vary between different provinces, states, or countries. For the early stages of the outbreak in the United States, in n = 50 states, we adopted the disease-specific values from China, and found a contact period of 3.38±0.69 days. Our network model predicts that–without the massive political mitigation strategies that are in place today–the United states would have faced a basic reproduction number of 5.3±0.95 and a nationwide peak of the outbreak on May 10, 2020 with 3 million infections. Our results demonstrate how mathematical modeling can help estimate outbreak dynamics and provide decision guidelines for successful outbreak control. We anticipate that our model will become a valuable tool to estimate the potential of vaccination and quantify the effect of relaxing political measures including total lock down, shelter in place, and travel restrictions for low-risk subgroups of the population or for the population as a whole.

## 1 Motivation

In December 2019, a local outbreak of pneumonia of initially unknown cause was detected in Wuhan, a city of 11 million people in central China [24]. The cause of the disease was identified as the novel severe acute respiratory syndrome coronavirus 2, SARS-CoV-2 [16]. Infection with the virus can be asymptomatic or can result in a mild to severe symptomatic disease, coronavirus disease 2019 or COVID-19. The majority of COVID-19 cases result in mild symptoms including fever, cough, shortness of breath, and respiratory distress [20]. Severe complications arise when the disease progresses to viral pneumonia and multi-organ failure. The SARSCoV-2 virus can spread quickly, mainly during close contact, but also through small droplets from coughing or sneezing [32]. After the first four cases were reported on December 29, the outbreak quickly spread from Wuhan across all provinces of mainland China, and, in the following two months, across the entire world. On March 11, 2020, the World Health Organization acknowledged the alarming levels of spread and severity, and characterized the COVID-19 situation as a pandemic [31]. As of today, April 4, 2020, COVID19 has affected 203 countries with a total of 1,201,483 reported cases, 64,690 deaths, and 264,467 recovered cases [9].

Figure 1 illustrates a typical timeline of COVID-19 in a single person and shows how this timeline maps onto an entire population. For this example [17], at day 0, a number of susceptible individuals are exposed to the virus and transition from the susceptible to the exposed state. Around at day 3, the exposed individuals become infectious. During this time, they can infect others, while not showing any symptoms themselves. The infectious period lasts for approximately 10 days. Around day 5, infectious individuals become symptomatic. This implies that they have potentially spread the disease for two days without knowing it. In the majority of (1 − *ν*_h_) of the population, the symptomatic period lasts for approximately 9 days. Around day 9, a severely affected population of *ν*_h_ are hospitalized and their hospitalization lasts for approximately 14 days. Around day 10, *ν*_c_ of the hospitalized population experiences critical conditions that last for approximately 10 days and end in (1 − *ν*_d_) of recovery and *ν*_d_ of death. For a hospitalization fraction of *ν*_h_ = 0.045, a critical conditions fraction of *ν*_c_ = 0.25, and a death fraction of *ν*_d_ = 0.50, 99.44% of the population recover and 0.56% die [17].

**Fig. 1.**
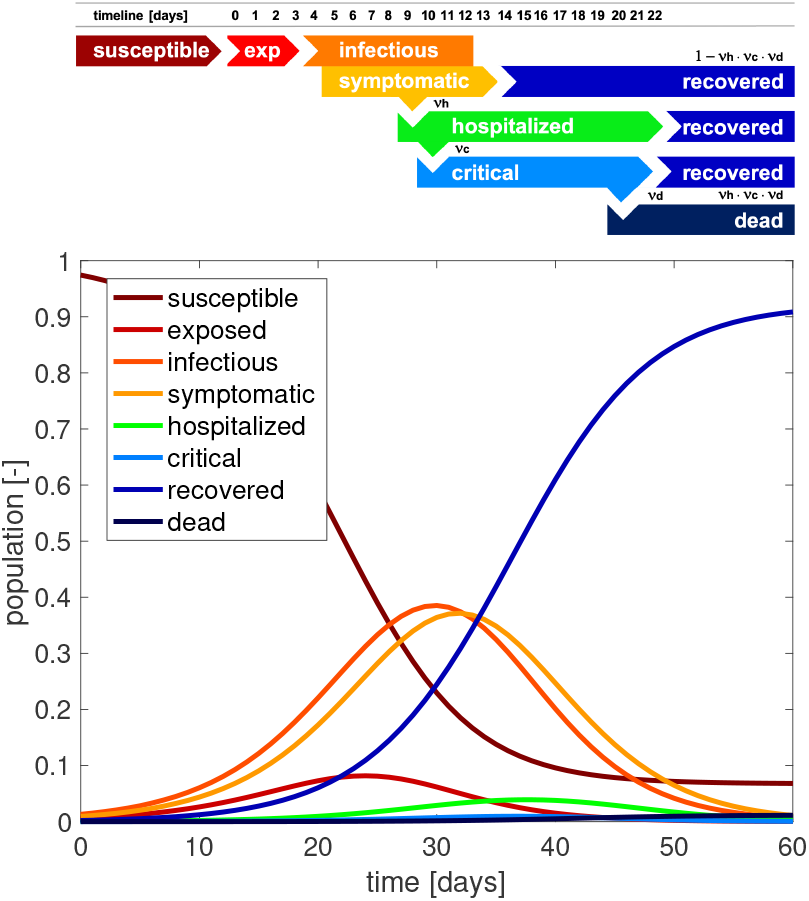
Typical timeline of COVID-19. At day 0, a fraction of the susceptible population is exposed to the virus. At day 3, exposed individuals become infectious, and the infectious period lasts for 10 days. At day 5, infectious individuals become symptomatic; the majority of the symptomatic population recovers after 9 days. At day 9, a fraction of the symptomatic population is hospitalized; the majority of the hospitalized population recovers after 14 days. At day 10, a fraction of the hospitalized population experiences critical conditions that last for 10 days and end in either recovery or in death. On the population level, the outbreak of COVID-19 can be summarized in eight curves that illustrate the dynamics of the individual subgroups.

The first mathematical models for infectious diseases date back to a smallpox model by Daniel Bernoulli in 1760 [4]. Since the 1920s, compartment models have become the most common approach to model the epidemiology of infectious diseases [21]. One of the simplest compartment models is the SEIR model that represents the timeline of a disease through four compartments, the susceptible, exposed, infectious, and recovered populations [2]. The temporal evolution of these compartments is governed by a set of ordinary differential equations parameterized in terms of the transition rates between them [18]. The transition rates *α* from the exposed to the infectious state and *γ* from the infectious to the recovered state are disease specific parameters. In fact, they are the inverses of the latent period *A* = 1/*α*, the time during which an individual is exposed but not yet infectious, and the infectious period *C* = 1/*γ*, the time during which an individual can infect others. This suggests that these two parameters are relatively independent of country, region, or city. In the example of Figure 1, the latent and infectious periods are *A* = 3 days and *C* = 10 days [17]. The most critical feature of the model is the transition from the susceptible to the exposed state. This transition is typically assumed to scale with the susceptible population *S*, the infectious population *I*, and the contact rate *β*, the inverse of the contact period *B* = 1/*β*, between them [23].

The product of the contact rate and the infectious period defines the basic reproduction number, *R*_0_ = *βC* = *C*/*B*, the number of individuals that are infected by a single one individual in an otherwise uninfected, susceptible population [11]. The basic reproduction number is a measure of the contagiousness or transmissibility of an infectious agent and it can vary considerably between different infectious diseases [10]. Typical basic reproduction numbers are on the order of 18 for measles, 9 for chickenpox, 7 for mumps, 7 for rubella, and 5 for poliomyelitis [1]. When the basic reproduction number is larger than one, *R*_0_ > 1.0, the infectious period *C* is larger than the contact period *B* [23]. This implies that at onset of an epidemic outbreak, when the entire population is susceptible, an infected individual will infect more than one other individual. In agreement with Figure 1, the infectious population first increases, then reaches a peak, and decreases toward zero [21]. As more and more individuals transition from the susceptible through the exposed and infectious states into the recovered state, the exposed and infectious states into the recovered state, the susceptible populations decreases. Once a large enough fraction of a population has become immune–either through recovery from the infection or through vaccination–this group provides a measure of protection for the susceptible population and the epidemic dies out [11]. This indirect protection is called herd immunity [14]. The concept of herd immunity implies that the converged susceptible population at endemic equilibrium is always larger than zero, *S*_∞_ > 0, and its value depends on the basic reproduction number *R*_0_. For a given basic reproduction number *R*_0_, herd immunity occurs at an immune fraction of (1 − 1/*R*_0_). Knowing the basic reproduction number is therefore critical to estimate the immune fraction of the population that is required to eradicate an infectious disease, for example, 94.4% for measles and 80.0% for poliomyelitis [18].

Restrictive measures like medical isolation or quarantine reduce the effective infectious period C and mitigation strategies like contact tracing, physical distancing, or travel restrictions increase the contact period B. Especially during the early stages of an outbreak, passenger air travel can play a critical role in spreading a disease [3], since traveling individuals naturally have a disproportionally high contact rate [30]. Border control can play a pivotal role in mitigating epidemics and prevent the spreading between cities, states, or countries [34]. In an attempt to mitigate the COVID-19 outbreak, many countries have implemented travel restrictions and mandatory quarantines, closed borders, and prohibited non-citizens from entry. This has stimulated an ongoing debate about how strong these restrictions should be and when it would be safe to lift them. The basic reproduction number is *R*_0_ provides guidelines about the required strength of political counter measures [18]. However, empirically finding the basic reproduction number requires careful contact tracing and is a lot of work, especially once the number of infectious individuals has grown beyond an overseeable size [24]. Network modeling of travel-induced spreading can play an important role in estimating the value of *R*_0_ [7] and interpreting the impact of travel restriction and border control [19].

## 2 Methods

### 2.1 Epidemiology modelling

We model the epidemiology of the COVID-19 outbreak using an SEIR model with four compartments, the susceptible, exposed, infectious, and recovered populations, governed by a set of ordinary differential equations [18],

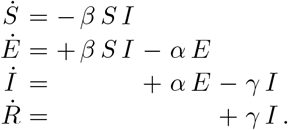

The transition rates between the four compartments, *β, α*, and *γ* are inverses of the contact period *B* = 1/*β*, the latent period *A* = 1/*α*, and the infectious period *C* = 1/*γ*. We interpret the latent and infectious periods *A* and *C* as disease-specific, and the contact period *B* as behavior specific. We discretize the SEIR model in time using an implicit Euler backward scheme and adopt a Newton Raphson method to solve for the daily increments in each compartment.

### 2.2 Network modeling

We model the spreading of COVID-19 across a country through a network of passenger air travel, which we represent as a weighted undirected graph 𝒢 with *N* nodes and *E* edges. The nodes represent the individual states, the edges the connections between them. We weight the edges by the estimated annual incoming and outgoing passenger air travel as reported by the Bureau of Transportation Statistics [6]. We summarize the connectivity of the graph 𝒢 in terms of the adjacency matrix *A*_*IJ*_, the frequency of travel between two states *I* and J, and the degree matrix *D*_*II*_ = diag 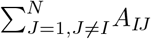, the number of incoming and outgoing connections of state I. The difference between the degree matrix *D*_*IJ*_ and the adjacency matrix *A*_*IJ*_ defines the weighted graph Laplacian *L*_*IJ*_,

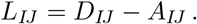

Figure 2 illustrates the discrete graph 𝒢 of the United States with *N* = 50 nodes and the *E* = 200 most travelled edges. The size and color of the nodes represent the degree DII, the thickness of the edges represents the adjacency *A*_*IJ*_. For our passenger travel-weighted graph, the degree ranges from 100 million in California to less than 1 million in Delaware, Vermont, West Virginia, and Wyoming, with a mean degree of 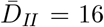 million per node. We assume that the Laplacian *L*_*IJ*_, normalized to one and scaled by the travel coefficient *ϑ*, characterizes the global spreading of COVID-19 and discretize our SEIR model on our weighted graph 𝒢. Specifically, we introduce the susceptible, exposed, infectious, and recovered populations *S*_*I*_, *E*_*I*_, *I*_*I*_, and *RI* as global unknowns at the *I* = 1, …, *N* nodes of the graph 𝒢. This results in the spatial discretization of the set of equations with 4 *N* unknowns,

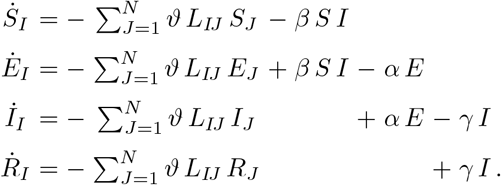

**Fig. 2.**
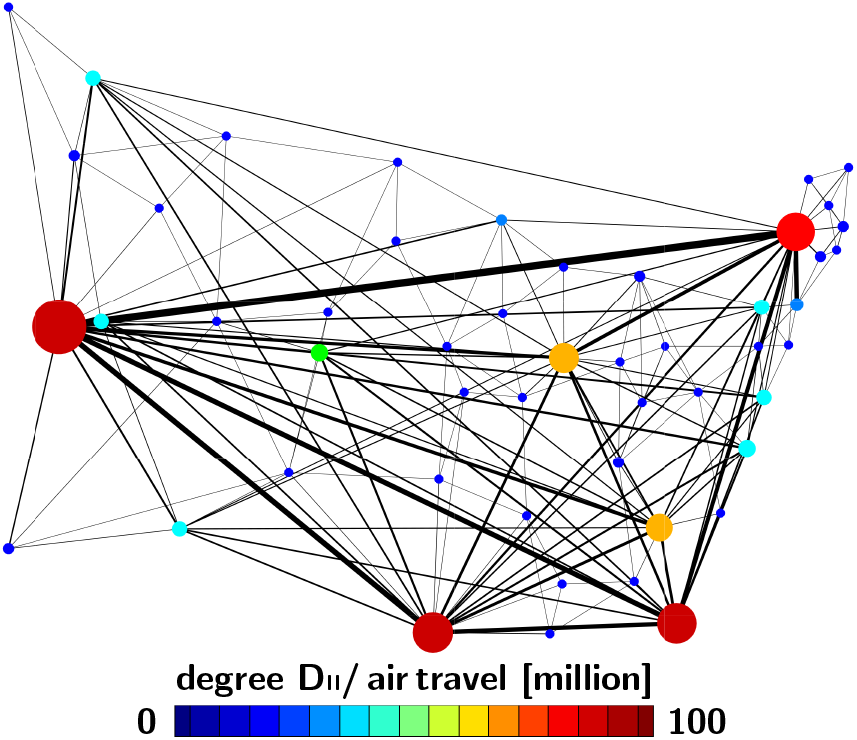
Network model of COVID-19 spreading across the United States. Discrete graph 𝒢 of the United States with *N* = 50 nodes and the 200 most travelled edges. Size and color of the nodes represent the degree *D*_*II*_, thickness of the edges represents the adjacency *A*_*IJ*_ estimated from annual incoming and outgoing passenger air travel.

We discretize our SEIR network model in time using an implicit Euler backward scheme and adopt a Newton Raphson method to solve for the daily increments in each compartment in each state [15].

### 2.3 Parameter identification

#### 2.3.1 COVID-19 outbreak dynamics in China

Unlike many other countries, China has already seen a peak of the COVID-19 outbreak and is currently not seeing a significant number of new cases. The COVID19 outbreak data of the Chinese provinces capture all three phases, increase, peak, and decrease of the infectious population and are currently the richest dataset available to date. This dataset describes the temporal evolution of confirmed, recovered, active, and death cases starting January 22, 2020 [9]. As of April 4, there were 81,639 confirmed cases, 76,755 recovered, 1,558 active, and 3,326 deaths. From these data, we map out the temporal evolution of the infectious group *I* as the difference between the confirmed cases minus the recovered and deaths, and the recovered group *R* as the sum of the recovered and deaths in each Chinese province. To simulate the province-specific epidemiology of COVID19 with the SEIR model, we use these data to identify the latent period *A* = 1/*α*, the infectious period *C* = 1/*γ*, and the contact period *B* = 1/*β* as a direct measure of the basic reproduction number *R*_0_ = *B*/*C*. As our sensitivity analysis in Figure 3 shows, the dynamics of the SEIR model depend critically on the initial conditions, the number of susceptible *S*_0_, exposed *E*_0_, infectious *I*_0_, and recovered *R*_0_ individuals on the day the very first infectious case is reported, *I*_0_ ≥ 1. Naturally, on this day, the recovered population is *R*_0_ = 0. Since the exposed population is asymptomatic, its initial value *E*_0_ is unknown. To quantify the initial exposed population *E*_0_, we introduce a parameter *ρ* = *E*_0_/*I*_0_, the initial latent population [27]. It defines the fraction of exposed versus infectious individuals at day 0 and is a measure of initial hidden community spreading. The fraction of the initial susceptible population, *S*_0_ = 1 − *E*_0_ − *I*_0_ − *R*_0_, ensures that the total population sums up to one. To map the total population of one onto the absolute number of cases for each province, we introduce the normalization parameter *η* = *N*^∗^/*N*, the affected population. It defines the fraction of the province-specific epidemic subpopulation *N*^∗^ relative to the province population *N* [29]. Altogether we identify five parameters for each province, the exposed period *A* = 1/*α*, the infectious period *C* = 1/*γ*, the contact period *B* = 1/*β* or the basic reproduction number *R* = *C*/*B*, the initial latent population *ρ* = *E*_0_/*I*_0_, and the affected population *η* = *N*^∗^/*N*. We performed the parameter identification using the Levenberg-Marquardt method of least squares. In this identification process, we ignored data from secondary outbreaks [9].

**Fig. 3.**
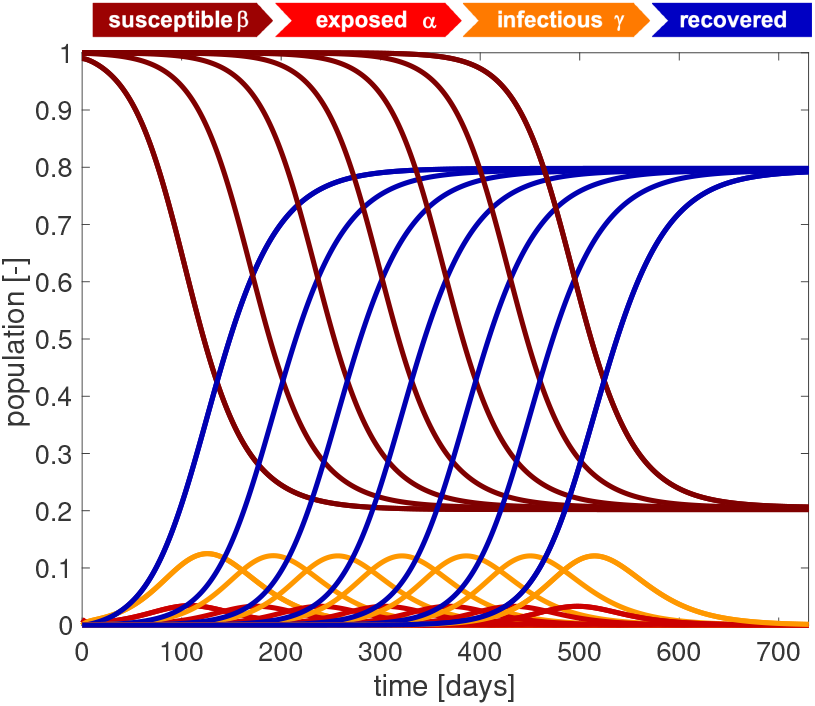
Outbreak dynamics. Sensitivity with respect to the initial exposed population *E*_0_. Decreasing the initial exposed population delays the onset of the outbreak while the shapes of all four curves remain the same. The susceptible and recovered populations converge to the same endemic equilibrium at *S*_∞_ = 0.202 and *R*_∞_ = 0.798. For an initial exposed population of *E*_0_ = 0.01, the infectious population reaches its maximum at *I*_max_ = 0.121 after 125 days. Decreasing the initial exposed population by a factor 10 delays the maximum by 65 days. Latent period *A* = 5 days, infectious period *C* = 20 days, basic reproduction number *R*_0_ = *C*/*B* = 2.0, and initial exposed population *E*_0_ = 10^−2^, 10^−3^, 10^−4^, 10^−5^, 10^−6^, 10^−7^, 10^−8^.

#### 2.3.2 COVID-19 outbreak dynamics in the United States

Unlike China, the United States are at the early stage of the COVID-19 outbreak and all states are still seeing an increase of the number of new cases every day. The available dataset describes the temporal evolution of confirmed, recovered, active, and death cases starting January 21, 2020, the first day of the outbreak in the United States [9]. As of April 4, there were 311,357 confirmed cases, 14,825 recovered, 288,081 active, and 8,451 deaths. Similar to the Chinese data, we map out the temporal evolution of the infectious group *I* as the difference between the confirmed cases minus the recovered and deaths in each state state of the United States. To simulate the state-specific epidemiology of COVID19 with the SEIR model, we use these data to identify the contact time *B* = 1/*β*, while fixing the disease-specific latent and infections periods *A* = 1/*α* and *C* = 1/*γ* at their mean values of the SEIR dynamics fit for the Chinese provinces, and indirectly fitting the basic reproduction number *R*_0_ = *C*/*B*. For each state, we set the first day of reported infections *I*_0_ ≥ 1 to day zero, at which the recovered population is *R*_0_ = 0, the unknown exposed population is *E*_0_ = *ρ I*_0_ [27], and the susceptible population is *S*_0_ = *N* − *E*_0_− *I*_0_− *R*_0_, where *N* is the state-specific population [33]. We identify two parameters for each state, the contact period *B* = 1/*β* and the initial latent population *ρ* = *E*_0_/*I*_0_, while we use the exposed period *A* = 1/*α* and the infectious period *C* = 1/*γ* from the parameter identification for the Chinese provinces and back-calculate the basic reproduction number R = *C*/*B*. We perform the parameter identification using the Levenberg-Marquardt method of least squares.

## 3 Results

### 3.1 Outbreak dynamics

The dynamics of the SEIR model are determined by three parameters, the latent period *A* = 1/*α*, and the infectious period *C* = 1/*γ*, and the contact period *B* = 1/*β*, or, alternatively, the basic reproduction number *R*_0_ = *C*/*B*. Before identifying these parameters for the outbreaks in China and in the United States, we will illustrate their effects by systematically varying each parameter while keeping the other values fixed. Specifically, unless stated otherwise, we choose a latent period of *A* = 5 days, an infectious period of *C* = 20 days, a basic reproduction number of *R*_0_ = *C*/*B* = 2.0, and an initial exposed population *E*_0_ = 0.010.

Figure 3 illustrates the sensitivity of the SEIR model with respect to the size of the initial exposed population *E*_0_. Decreasing the initial exposed population from *E*_0_ = 10^−2^, 10^−3^, 10^−4^, 10^−5^, 10^−6^, 10^−7^, 10^−8^ delays the onset of the outbreak while the dynamics of the susceptible, exposed, infectious, and recovered populations remain the same. For all seven cases, the susceptible and recovered populations converge to the same endemic equilibrium with *S*_∞_ = 0.202 and *R*_∞_ = 0.798. The infectious population increases gradually, reaches its maximum at *I*_max_ = 0.121, and then decreases. For the largest initial exposed population of *E*_0_ = 0.01 this maximum occurs after 125 days. Decreasing the initial exposed population by a factor 10 delays the maximum by 65 days. This highlights the exponential nature of the model, which causes a constant delay for a logarithmic decrease of the exponential population, while the overall outbreak dynamics remain the same. In view of the COVID-19 outbreak, this supports the general notion that even a single individual can cause an outbreak. If multiple individuals trigger the outbreak in a province, state, or country, the overall outbreak dynamics will remain the same, but the peak of the outbreak will happen earlier.

Figure 4 illustrates the sensitivity of the SEIR model with respect to the latent period A. Increasing the latent period from *A* = 0, 5, 10, 15, 20, 25 days increases the exposed population and decreases the infectious population. The susceptible and recovered populations converge to the same endemic equilibrium at *S*_∞_ = 0.202 and *R*_∞_ = 0.798. Convergence is slower for increased latent periods *A*. The steepest susceptible, infectious, and recovery curves correspond to the special case of the SIR model without a separate exposed population E, for which *A* = 0 days. This model does not have a separate exposed population. It reaches its peak infectious population of *I*_max_ = 0.157 after 86 days. In view of the COVID-19 outbreak this implies that knowledge of the latent period is important to correctly estimate the timing and peak of the infectious population, which ultimately determines the absolute number of hospital beds and ventilator units required to insure appropriate medical care.

**Fig. 4.**
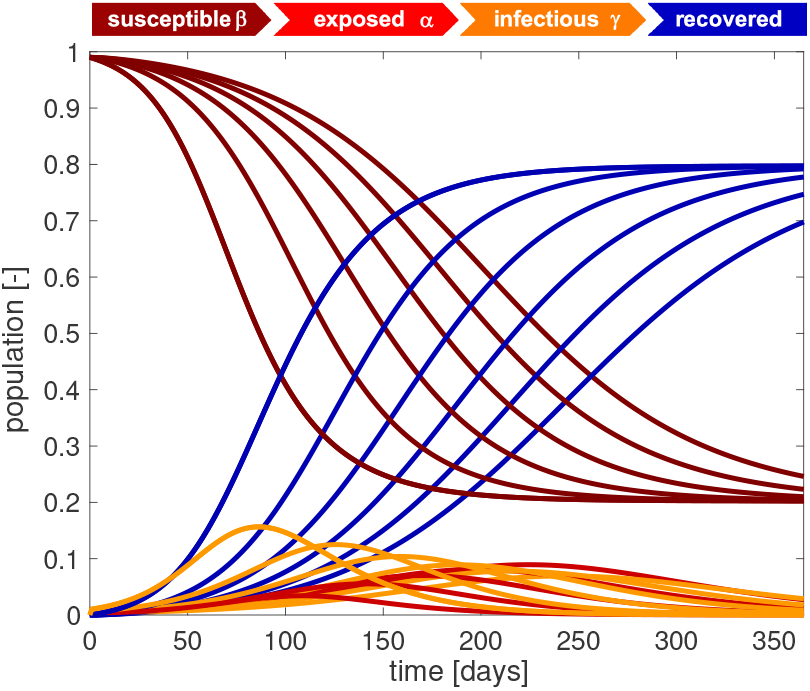
Outbreak dynamics. Sensitivity with respect to the latent period *A*. Increasing the latent period increases the exposed population and decreases the infectious population. The susceptible and recovered populations converge to the same endemic equilibrium at *S*_∞_ = 0.202 and *R*_∞_ = 0.798, however, slower. The steepest susceptible, infectious, and recovery curves correspond to the SIR model without separate exposed population *E* with *A* = 0 days with a maximum infectious population of *I*_max_ = 0.157 after 86 days. Latent period *A* = 0, 5, 10, 15, 20, 25 days, infectious period *C* = 20 days, basic reproduction number *R*_0_ = *C*/*B* = 2.0, and initial exposed fraction *E*_0_ = 0.010.

Figure 5 illustrates the sensitivity of the SEIR model with respect to the infectious period *C*. Increasing the infectious period at a constant basic reproduction number flattens the exposed population and increases the infectious population. The susceptible and recovered populations converge to the same endemic equilibrium at *S*_∞_ = 0.202 and *R*_∞_ = 0.798, however, slower. The flattest susceptible, infectious, and recovery curves correspond to longest infectious period of *C* = 30 days and a contact period of *B* = 15 days with the maximum infectious population of *I*_max_ = 0.135 after 169 days. In view of the COVID-19 outbreak, knowing the infectious time is important to correctly estimate the timing and peak of the infectious population, and with days. In view of the COVID-19 outbreak, knowing the units.

**Fig. 5.**
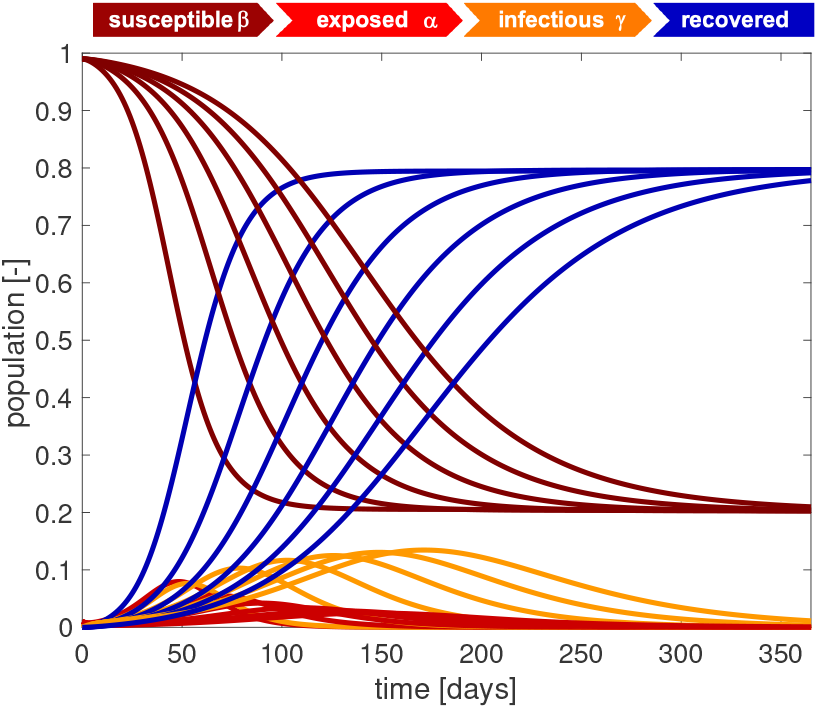
Outbreak dynamics. Sensitivity with respect to the infectious period *C*. Increasing the infectious period at a constant basic reproduction number flattens the exposed population and increases the infectious population. The susceptible and recovered populations converge to the same endemic equilibrium at *S*_∞_ = 0.202 and *R*_∞_ = 0.798, however, slower. The flattest susceptible, infectious, and recovery curves correspond to longest infectious period of *C* = 30 days with the maximum infectious population of *I*_max_ = 0.135 after 169 days. Latent period *A* = 5 days, infectious period *C* = 5, 10, 15, 20, 25, 30 days, basic reproduction number *R*_0_ = *C*/*B* = 2.0, and initial exposed fraction *E*_0_ = 0.010.

Figure 6 illustrates the sensitivity of the SEIR model with respect to the basic reproduction number *R*_0_. Decreasing the basic reproduction number decreases the exposed and infectious populations. The susceptible and recovered populations converge to larger and smaller endemic equilibrium values, and converges is slower. The steepest susceptible, exposed, infectious, and recovery curves correspond to the largest basic reproduction number of *R*_0_ = 10.0 with the maximum infectious population of *I*_max_ = 0.488 after 35 days and converge to an endemic equilibrium at *S*_∞_ = 0.0001 and *R*_∞_ = 0.9999. In view of the COVID-19 outbreak, the basic reproduction number is the parameter that we can influence by political counter measures. Reducing the basic reproduction number beyond its natural value by decreasing the contact time *B* through physical distancing or total lock down allows us to reduce the maximum infectious population and delay the outbreak, a measure that is commonly referred to in the public media as “flatting the curve”.

**Fig. 6.**
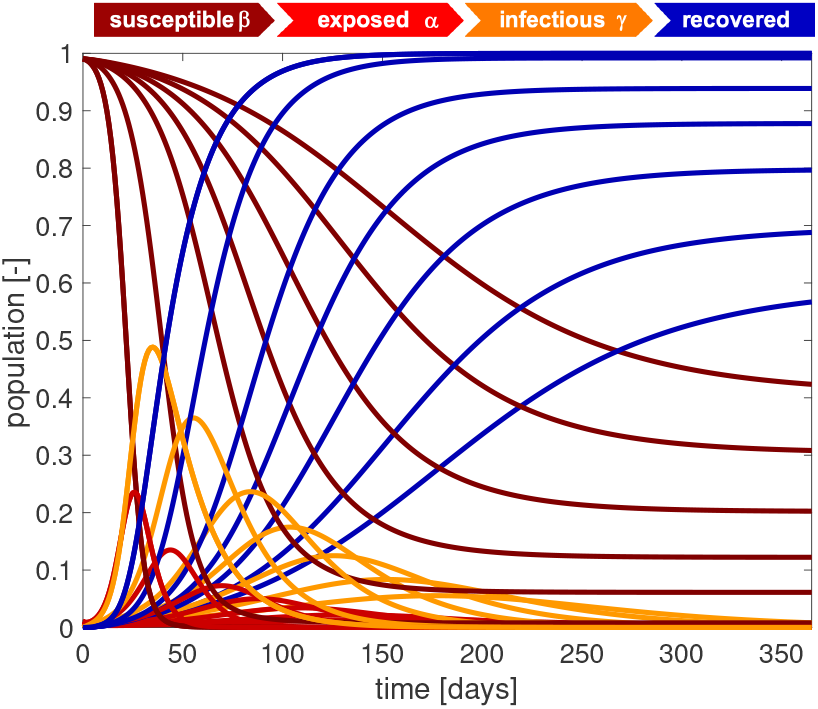
Outbreak dynamics. Sensitivity with respect to the basic reproduction number *R*_0_. Decreasing the basic reproduction number decreases the exposed and infectious populations. The susceptible and recovered populations converge to larger and smaller endemic equilibrium values, and converges is slower. The steepest susceptible, exposed, infectious, and recovery curves correspond to the largest basic reproduction number of *R*_0_ = 10.0 with the maximum infectious population of *I*_max_ = 0.488 after 35 days and converge to an endemic equilibrium at *S*_∞_ = 0.0001 and *R*_∞_ = 0.9999. Latent period *A* = 5 days, infectious period *C* = 20 days, basic reproduction number *R*_0_ = *C*/*B* = 1.5, 1.7, 2.0, 2.4, 3.0, 5.0, 10.0, and initial exposed fraction *E*_0_ = 0.010.

### 3.2 Outbreak control

The sensitivity study suggests that an epidemic outbreak is most sensitive to the basic reproduction number *R*_0_. While the latent period A and the infectious period *C* are disease specific, community mitigation and political action can modulate the basic reproduction number *R*_0_ through a variety of measures including active contact tracing, isolation of infectious individuals, quarantine of close contacts, travel restrictions, physical distancing, or total lock down.

Figure 7 illustrates the effect of the basic reproduction number *R*_0_ on the maximum exposed and infectious populations *E*_max_ and *I*_max_ and on the converged susceptible and recovered populations *S*_∞_ and *R*_∞_ at endemic equilibrium. Increasing the basic reproduction number beyond one increases the maximum exposed and infectious populations. The converged susceptible and recovered populations decrease towards zero and increase towards one. For the chosen latent and infectious periods of *A* = 5 days and *C* = 20 days, the time to reach the maximum infectious population reaches its maximum of 213 days at a basic reproduction number *R*_0_ = 1.22 and decreases for increasing basic reproduction numbers. In view of the COVID-19 outbreak, Figure 7 suggests strategies to modulate the timeline of the epidemic by reducing the basic reproduction number *R*_0_. For example, if we we have access to a certain number of intensive care unit beds and ventilators, and we know rates of the infectious population that have to be hospitalized and require intensive care, we need to limit the maximum size of the population that becomes infectious. To limit the infectious fraction to 20% of the total population, i.e., *I*_max_ = 0.200, we would have to reduce the basic reproduction number to *R*_0_ = 2.69. The gray line indicates that this maximum would occur after 0.25 years or 93 days.

**Fig. 7.**
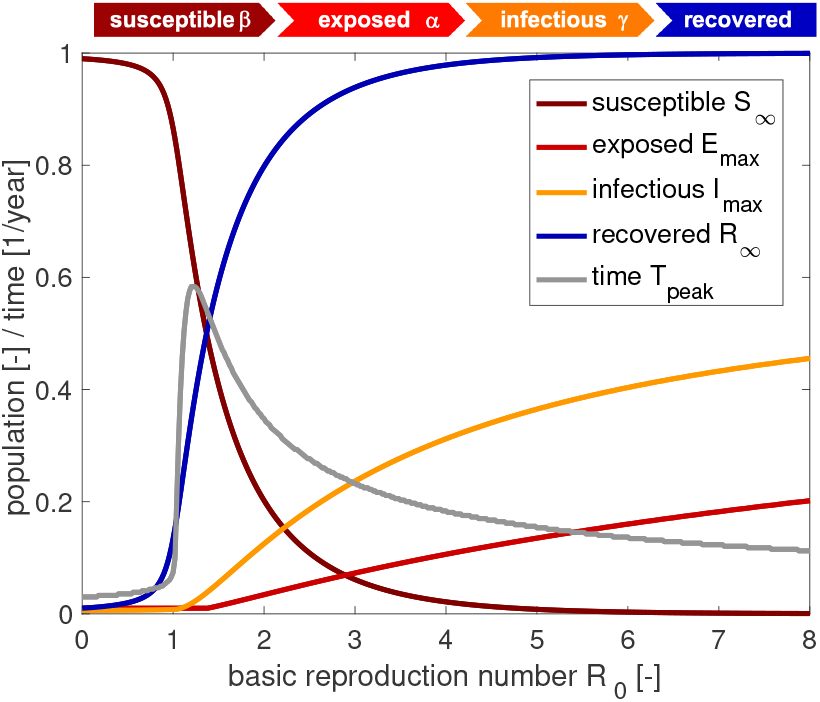
Outbreak control. Effect of basic reproduction number *R*_0_. Increasing the basic reproduction number beyond one increases the maximum exposed and infectious populations *E*_max_ and *I*_max_. The converged susceptible and recovered populations *S*_∞_ and *R*_∞_ at endemic equilibrium converge towards zero and one. The time to reach the maximum infectious population reaches its maximum of 213 days at a basic reproduction number *R*_0_ = 1.22 and decreases for increasing basic reproduction numbers. Latent period *A* = 5 days, infectious period *C* = 20 days, basic reproduction number, and initial exposed fraction *E*_0_ = 0.010.

Figure 8 illustrates the effect of constraining the outbreak by increasing the basic reproduction number R(*t*) such that the infectious population always remains below a tolerated infectious population, *I* < *I*_tol_. Decreasing the tolerated infectious population, *I*_tol_ = 0.15, 0.10., 0.08, 0.06, 0.05, 0.04, 0.03, 0.02 0.02, increases the required level of containment and decreases the relative basic reproduction number, *R*_0_(*t*)/*R*_0_ = 1.000, 0.742, 0.661, 0.603, 0.580, 0.541, 0.535, 0.524. This has the desired effect of decreasing the exposed and infectious populations. The susceptible population converges to progressively larger endemic equilibrium values *S*_∞_ =0.202, 0.225, 0.248, 0.274, 0.290, 0.309, 0.331, 0.358. The recovered population converges to progressively smaller endemic equilibrium values *R*_∞_ =0.798, 0.775, 0.752, 0.726, 0.710, 0,691, 0.669, 0.642. Convergens is slower under constrained outbreak. The lowest exposed and infectious curves and the flattest susceptible and recovery curves correspond to the most constrained infectious population of *I*_tol_ = 0.02 with a required level of containment of *R*_0_(*t*)/*R*_0_ = 0.524. The highest exposed and infectious curves and the steepest susceptible and recovery curves correspond to an unconstrained infectious population *I*_tol_ = 0.150 > *I*_max_ = 0.121 with peak infection after 125 days. In view of the COVID-19 outbreak, the gray line tells us how drastic political counter measures need to be. A required level of containment of *R*_0_(*t*)/*R*_0_ = 0.524 implies that we need to reduce the number of infections of a single individual by about one half. However, reducing the maximum infectious population comes at a socioeconomic price: The graphs teach us that it is possible to reach an endemic equilibrium at a smaller total number of individuals that have had the disease; yet, this endemic equilibrium would occur much later in time, for this example, after two or three years.

**Fig. 8.**
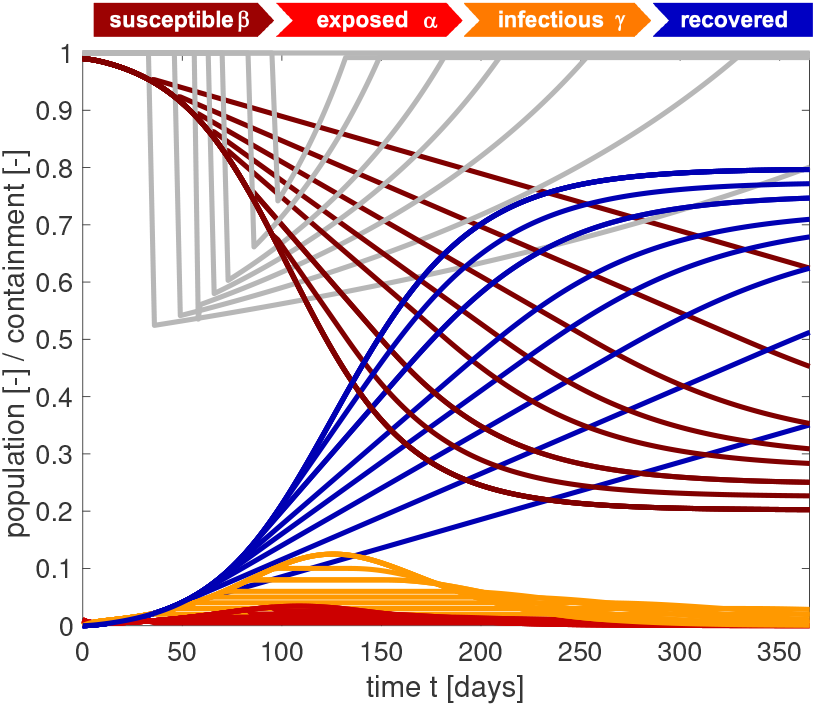
Outbreak control. Sensitivity with respect to tolerated infectious population *I*_tol_. Decreasing the tolerated infectious population increases the required level of containment *R*_0_(*t*)/*R*_0_. This decreases the exposed and infectious populations. The susceptible and recovered populations converge to larger and smaller endemic equilibrium values, but their converges is slower. tolerated infected population *I*_tol_ = 0.02, 0.03, 0.04, 0.05, 0.06, 0.08, 0.10, 0.15, basic reproduction number *R*_0_(*t*), and initial exposed fraction *E*_0_ = 0.010.

**Fig. 9.**
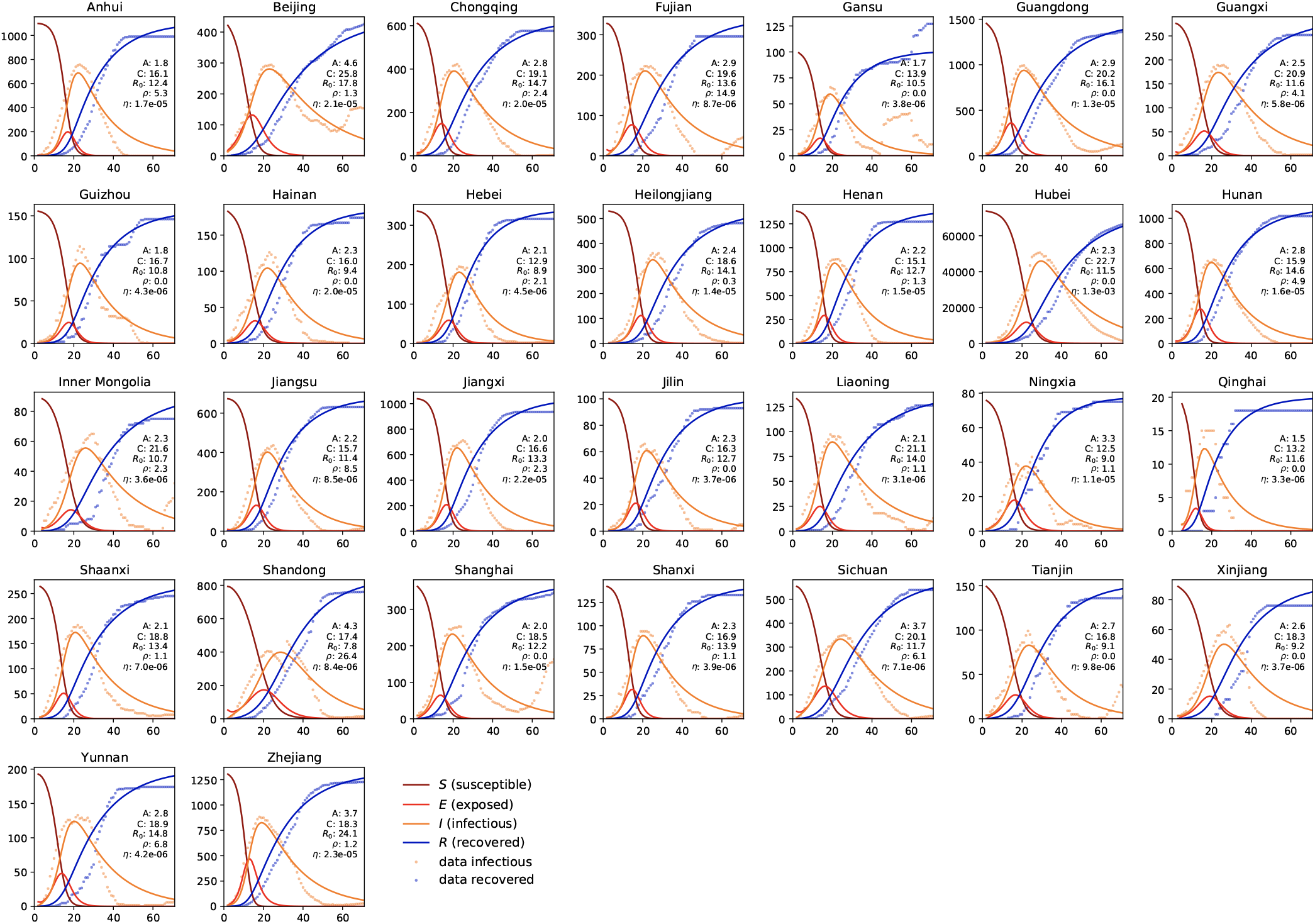
COVID-19 outbreak dynamics in China. Reported infectious and recovered populations and simulated susceptible, exposed, infectious, and recovered populations. Simulations are based on a province-specific parameter identification of the latent period *A*, contact period *B*, and infectious period *C*, defining the basic reproduction number *R*_0_ = *C*/*B*, the fraction of the initial latent population *ρ* = *E*_0_/*I*_0_, and the fraction of the affected population *η* = *N*^∗^/*N* for each province.

### 3.3 COVID-19 outbreak dynamics in China

Figure 9 summarizes the dynamics of the COVID-19 outbreak in 30 Chinese provinces. The dots indicate the reported infectious and recovered populations, the lines highlight the simulated susceptible, exposed, infectious, and recovered populations. The simulations are based on a province-specific parameter identification of the latent period A, the contact period B, the infectious period C, and from both, the basic reproduction number *R*_0_ = *C*/*B*, the fraction of the initial latent population *ρ* = *E*_0_/*I*_0_, and the fraction of the affected population *η* = *N*^∗^/*N* for each province. These five province-specific values are reported in each graph. Notably, the province of Hubei, where the outbreak started, has seen the most significant impact with more than 60,000 cases. Naturally, in Hubei, where the first cases were reported, the fraction of the initial latent population *ρ* is zero. Small values of *ρ* indicate a close monitoring of the COVID-19 outbreak, with very few undetected cases at the reporting of the first infectious case. The largest value of *ρ* = 26.4 suggests that, at the onset of the outbreak, a relatively large number of cases in the province of Shandong was undetected. The fraction of the affected population *η* = *N*^∗^/*N* is a province-specific measure for the containment of the outbreak. Naturally, this number is largest in the province of Hubei, with *η* = 1.3 · 10^−3^, and, because of strict containment, much smaller in all other provinces.

Table 1 summarizes the parameters for the COVID-19 outbreak in China. Averaged over all Chinese provinces, we found a latent period of *A* = 2.56±0.72 days, a contact period of *B* = 1.47±0.32 days, an infectious period of *C* = 17.82 ± 2.95 days, a basic reproduction number of *R*_0_ = *C*/*B* = 12.58±3.17, a fraction of the initial latent population of *ρ* = *E*_0_/*I*_0_ = 3.19±5.44, and fraction of the affected population of *η* = *N*^∗^/*N* = 5.19·10−5± 2.23·10−4.

**Table 1.**
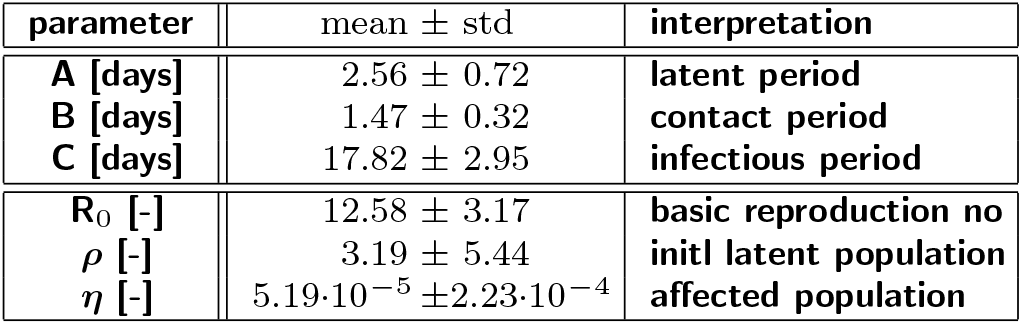
COVID-19 outbreak dynamics in China. Latent period *A*, contact period *B*, infectious period *C*, basic reproduction number *R*_0_ = *C*/*B*, fraction of initial latent population *ρ* = *E*_0_/*I*_0_ and fraction of affected population *η* = *N*^∗^/*N*.

| parameter | mean ± std | interpretation |
| --- | --- | --- |
| **A [days]** | $2.56 \pm 0.72$ | **latent period** |
| **B [days]** | $1.47 \pm 0.32$ | **contact period** |
| **C [days]** | $17.82 \pm 2.95$ | **infectious period** |
| **$R_0$ [-]** | $12.58 \pm 3.17$ | **basic reproduction no** |
| **$\rho$ [-]** | $3.19 \pm 5.44$ | **initl latent population** |
| **$\eta$ [-]** | $5.19 \cdot 10^{-5} \pm 2.23 \cdot 10^{-4}$ | **affected population** |

### 3.4 COVID-19 outbreak dynamics in the United States

Figure 10 shows the dynamics of the early stages of the COVID-19 outbreak in the 50 states of the United States, the District of Columbia, and the territories of Guam, Puerto Rico, and the Virgin Islands. The dots indicate the reported cases and death, the lines highlight the simulated susceptible, exposed, infectious, and recovered populations. The simulations are based on a state-specific parameter identification of the contact period *B* that defines the basic reproduction number *R*_0_ = *C*/*B* and of the fraction of the initial latent population *ρ* = *E*_0_/*I*_0_ at a given outbreak delay d0 for each state. These three state-specific values are reported in each graph. Since the outbreak is currently still in its early stages, we do not attempt to identify the latent and infectious periods, but rather adopt the mean latent and infectious periods *A* = 2.56 and *C* = 17.82 from the Chinese outbreak in Table 1. Notably, the state of New York is currently seeing the most significant impact with more than 100,000 cases. Naturally, in Washington, Illinois, California, and Arizona where the first cases were reported, the fraction of the initial latent population *ρ* is small. Largest *ρ* values occur in New York, New Jersey, Michigan, and Louisiana. The largest basic reproduction numbers *R*_0_ are identified in Idaho, Puerto Rico, Pennsylvania, and Indiana.

**Fig. 10.**
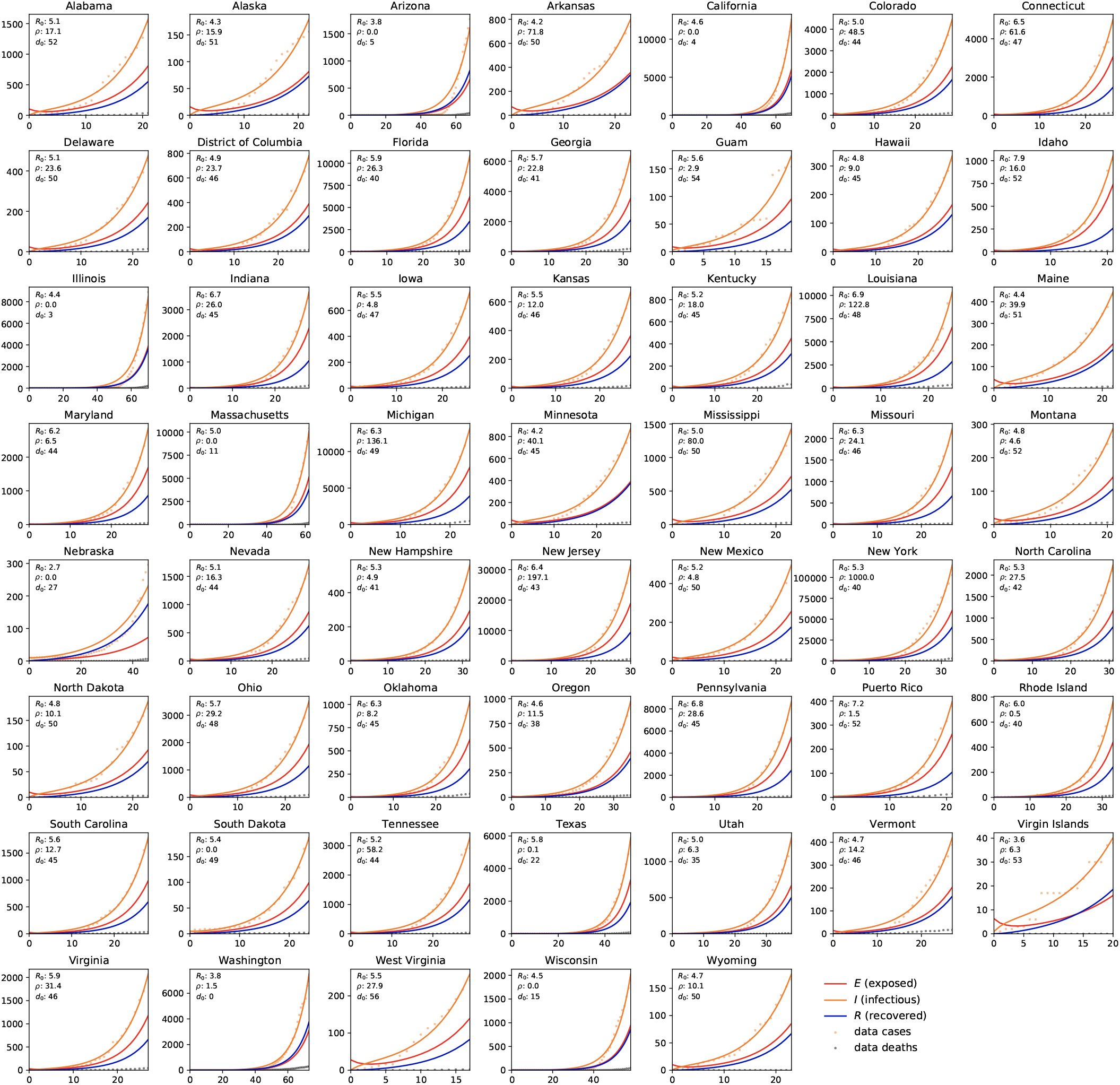
COVID-19 outbreak dynamics in the United States. Reported infectious populations and simulated exposed, infectious, and recovered populations. Simulations are based on a state-specific parameter identification of the contact period *B* defining the basic reproduction number *R*_0_ = *C*/*B*, and the fraction of the initial latent population *ρ* = *E*_0_/*I*_0_ for each state, for a given outbreak delay d_0_ and disease specific latent and infectious periods *A* = 2.56 and *C* = 17.82 identified for the Chinese outbreak.

Table 2 summarizes the parameters for the early stages of the COVID-19 outbreak in the United States. Averaged over all states, we found a contact period of *B* = 3.38 ± 0.69 days resulting in a basic reproduction number of *R*_0_ = *C*/*B* = 5.30 ± 0.95, a fraction of the initial latent population of *ρ* = *E*_0_/*I*_0_ = 43.75 ± 126.34 and an outbreak delay of d_0_ = 41.28 ± 13.78 days.

**Table 2.**
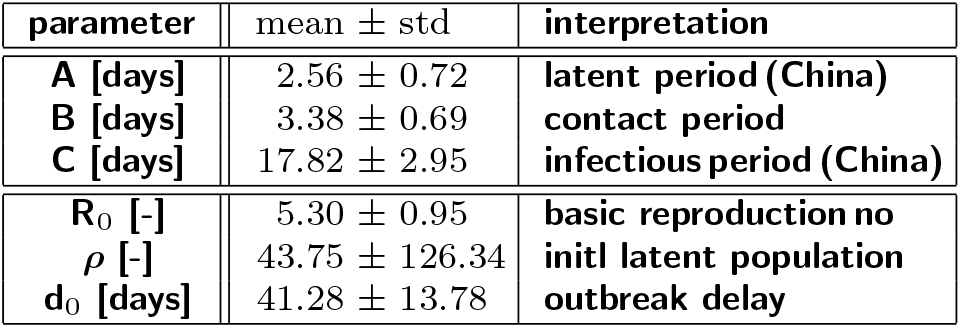
COVID-19 outbreak dynamics in the United States. Contact period *B*, basic reproduction number *R*_0_ = *C*/*B*, fraction of initial latent population *ρ* = *E*_0_/*I*_0_ and the outbreak delay using latent period *A* and infectious period *C* from the outbreak in China.

Figure 13 illustrates the undetected population at the onset of the outbreak across the United States. The *ρ* = *E*_0_/*I*_0_ value is small in the first states where the outbreak was reported, Washington, Illinois, California, and Arizona, suggesting that the reported cases were truly the first cases in those states. In states where the first cases occurred later, the *ρ* value increases. Notably, Louisiana, Michigan, New Jersey, and New York have the highest ρ values of 122.8, 136.1, 197.1, and 1,000 suggesting that both had an exceptionally high number of exposed individuals or individuals that were infected but unreported.

**Fig. 11.**
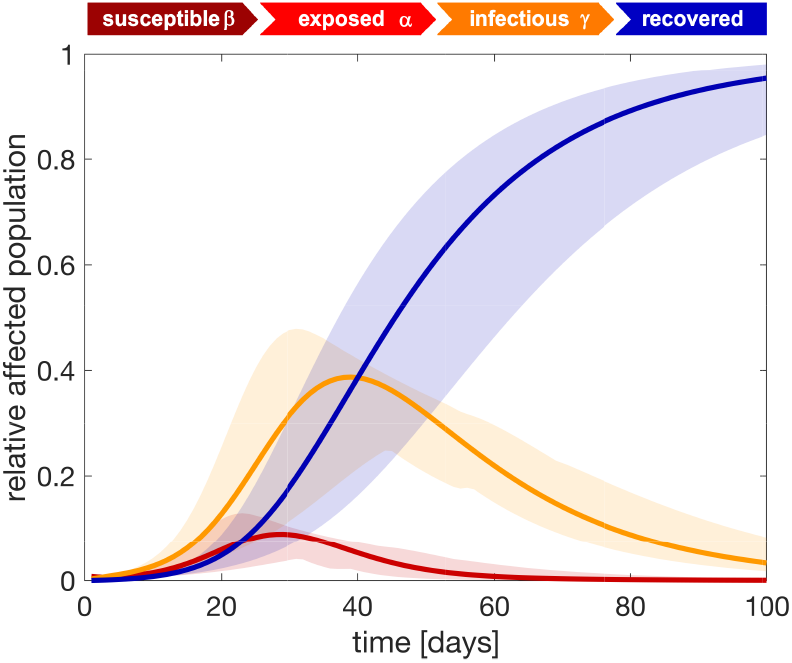
COVID-19 outbreak dynamics in the United States predicted with the SEIR model. Exposed, infectious, and recovered fractions of the affected populations for each state predicted using data from the early states of the outbreak and assuming no additional counter measures. Solid lines represent the mean and shaded regions highlight the 95% confidence interval. Latent period *A* = 2.56 days, contact period *B* = 3.38 days, infectious period *C* = 17.82 days, and fraction of initial latent population *ρ* = *E*_0_/*I*_0_ = 43.75.

**Fig. 12.**
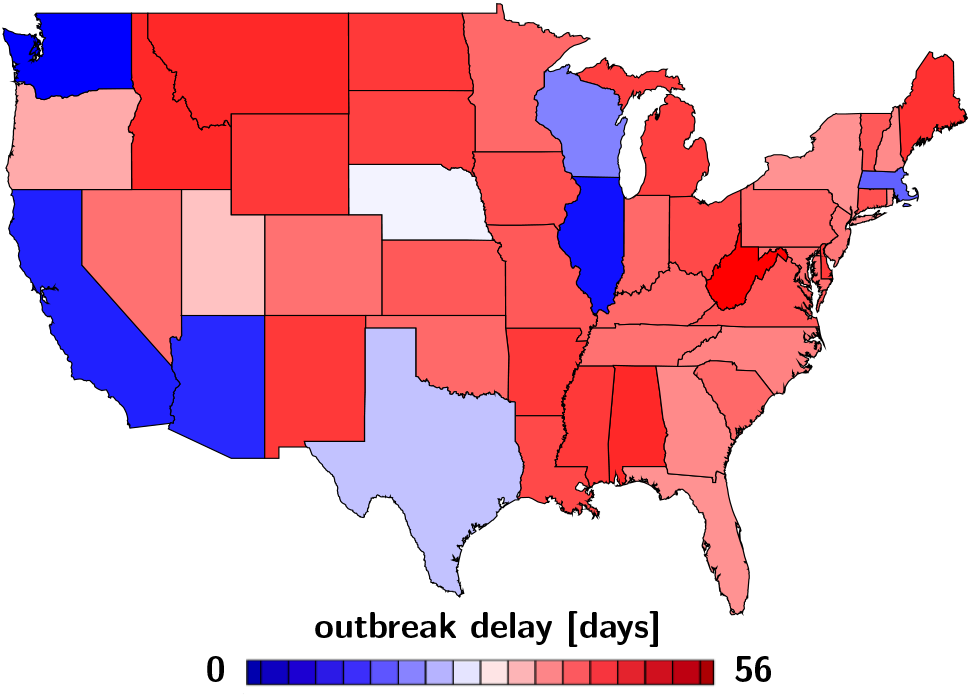
Regional variation of the outbreak delay d_0_. The outbreak varies from 0 days in Washington, the first state affected by the outbreak, to 56 days in West Virginia, the last state affected by the outbreak.

**Fig. 13.**
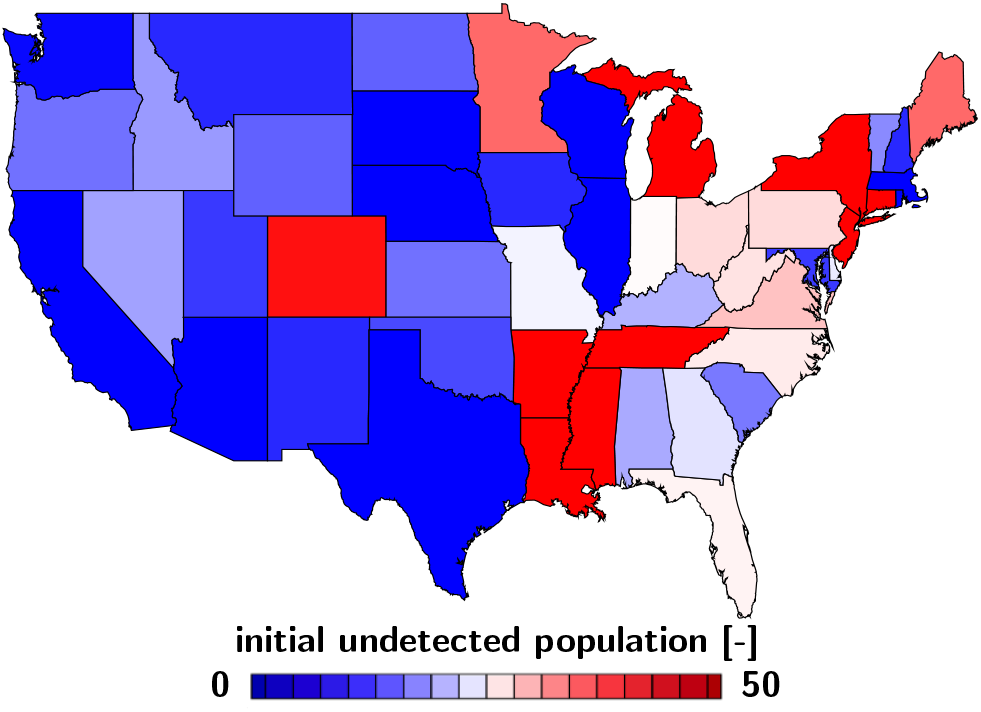
Regional variation of the initial undetected population *ρ*. The fraction of the initial undetected population is smallest in Washington, Illinois, California, and Arizona and largest in Louisiana with 122.8, Michigan with 136.1, New Jersey with 197.1, and New York with 1,000.

Figure 14 illustrates the basic reproduction number for the early stages of the outbreak across the United States. The basic reproduction number *R*_0_ = *C*/*B*, the number of individuals infected by a single infectious individual, varies from minimum values of 2.5 and 3.6 in Nebraska and Arizona to maximum values of 7.2 and 7.9 in Puerto Rico and Idaho.

**Fig. 14.**
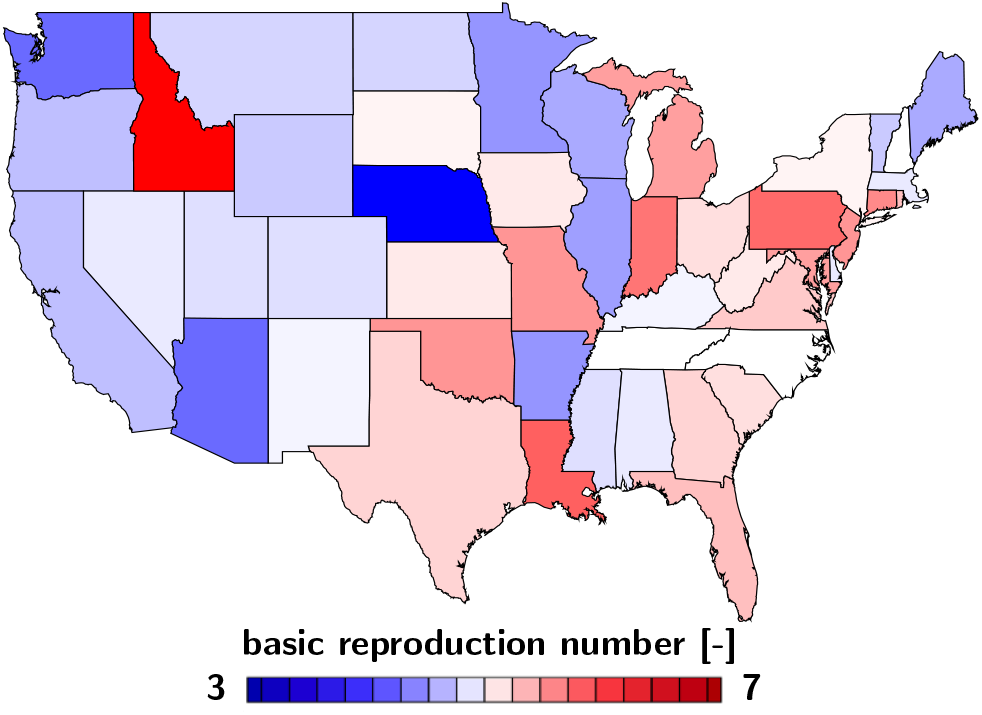
Reginal variation of the basic reproduction number R_0_. During the early stages of the outbreak, the basic reproduction number varies from minimum values of 2.5 and 3.6 in Nebraska and Arizona to maximum values of 7.2 and 7.9 in Puerto Rico and Idaho.

Figure 15 shows the nation-wide exposed, infectious, and recovered cases for the United States. The circles highlight the reported cases, the lines the predictions of the SEIR network model using data from the early stages of the outbreak with parameters from Tables 1 and 2 and a travel coefficient of *ϑ* = 0.43. The graphs starts on d_0_, the day at which the last state reported its first case d_0_ = March 17, 2020. Compared to the outbreak characteristics for the individual status in Figure 11 with a peak of the infectious population at 39 days after the first infectious case has been reported, the nation-wide outbreak peaks 54 days after the last state has been an outbreak, on May 10, 2020. This difference is a manifestation of both the state specific outbreak delay d_0_ and the travel of individuals between the different states represented through the network model. Figure 16 illustrates the spatio-temporal evolution of the infection population across the United States as predicted by the SEIR network model. The simulation is based on data from the early stages of the outbreak and assumes that no additional counter measures have been implemented. Days 10 and 20 illustrate the slow growth of the infectious population during the early states of the outbreak. The state of New York sees the outbreak first, followed by New Jersey and Louisiana. Days 30 and 40 illustrate how the outbreak spreads across the country. With no additional counter measures, the SEIR network model predicts a nation-wide peak of the outbreak on day 54, on May 10, 2020. Day 50 illustrates that the earlier affected states, New York, New Jersey, and Louisiana already see a decrease of the infected population. Nebraska, West Virginia, and Wisconsin are still far from reaching the peak. Compared to Figures 12 to 14 these maps account for both, the outbreak delay and the travel of individuals between the different states represented through the network model. This model would allow us to probe the effect of travel restrictions to and from a specific state by locally reducing its travel coefficients or by globally reducing the nation-wide transport coefficient across the United States.

**Fig. 15.**
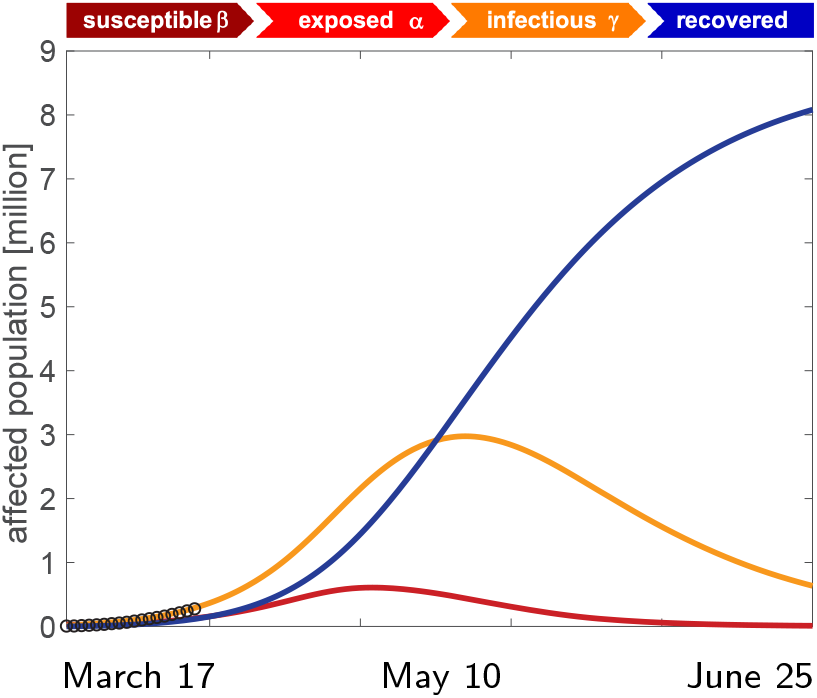
COVID-19 outbreak dynamics across the United States predicted with the SEIR network model. Exposed, infectious, and recovered cases for the United States reported and predicted by the SEIR network model using data from the early stages of the outbreak. With no additional counter measures, the SEIR network model predicts a nation-wide peak of the outbreak on day 54, on May 10, 2020. Latent period *A* = 2.56 days, contact period *B* = 3.38 days, infectious period *C* = 17.82 days, fraction of initial latent population *ρ* = *E*_0_/*I*_0_ = 43.75, day at which the last state reported its first case d_0_ = March 17, 2020, and travel coefficient *ϑ* = 0.43

**Fig. 16.**
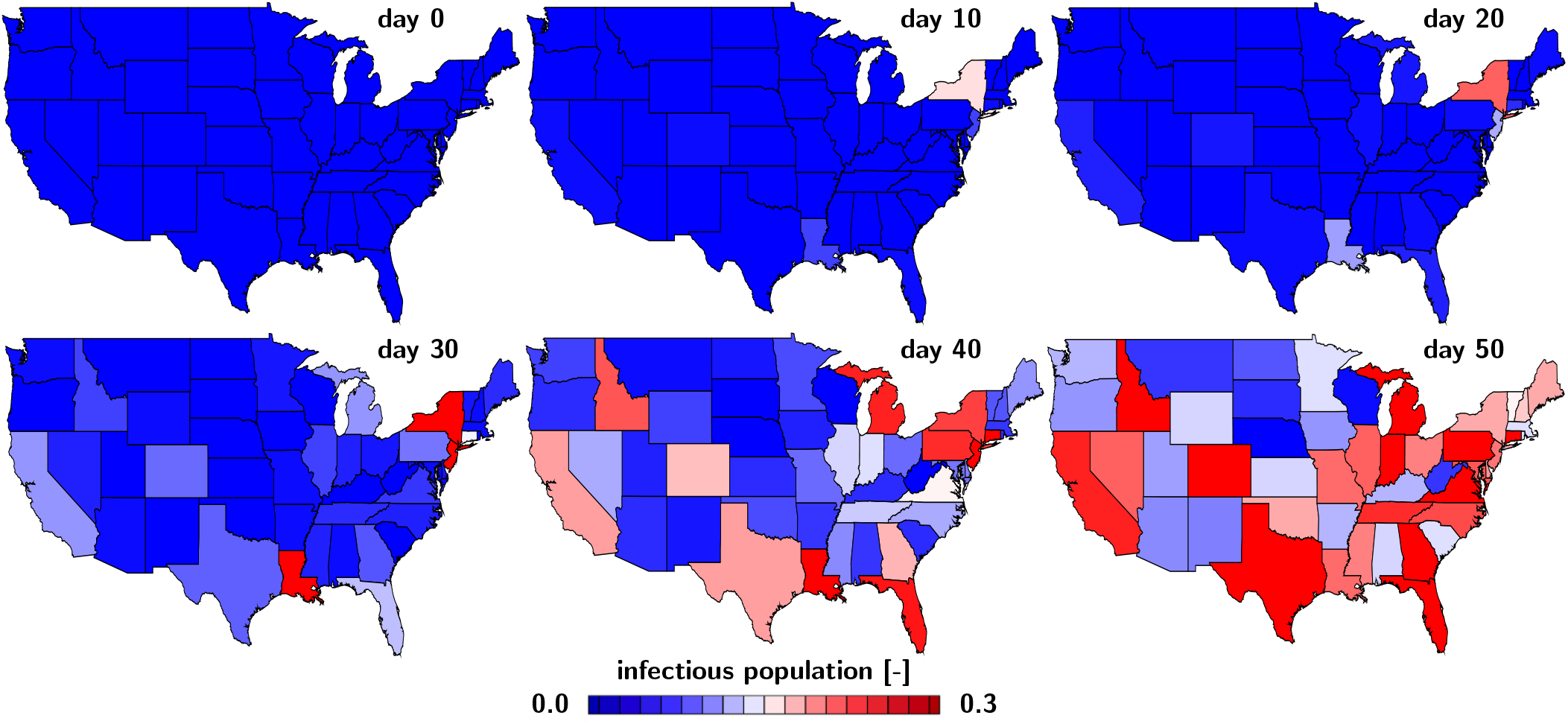
COVID-19 outbreak dynamics across the United States predicted with the SEIR network model. Regional evolution of the infectious population *I* predicted by the SEIR network model using data from the early stages of the outbreak. Days 10 and 20 illustrate the slow growth of the infectious population during the early stages of the outbreak. The state of New York sees the outbreak first, followed by New Jersey, Louisiana, and California. Days 30 and 40 illustrate how the outbreak spreads across the country. With no additional counter measures, the SEIR network model predicts a nation-wide peak of the outbreak on day 54, on May 10,2020. Day 50 illustrates that the earlier affected states, New York, New Jersey, and Louisiana already see a decrease of the infected population, while other states like Nebraska, West Virginia, and Wisconsin are still far from reaching the peak. Latent period *A* = 2.56 days, contact period *B* = 3.38 days, infectious period *C* = 17.83 days, fraction of initial latent population *ρ* = *E*_0_/*I*_0_ = 43.75, day at which the last state recorded an outbreak d_0_ = March 17, 2020, and travel coefficient *ϑ* = 0.43.

## 4. Discussion

We have established a simulation tool that can estimate the dynamics of the COVID-19 outbreak, both locally for individual provinces or states and globally for an entire country. Our simulations suggest that–despite the social, regional, demographical, geographical, and socio-economical heterogeneities in different regions– the outbreak of COVID-19 follows a universal model with a few relatively robust parameters. Specifically, our simulation integrates a global network model with a local epidemic SEIR model at each network node. It uses six epidemiologically meaningful parameters, the latent and infectious periods *A* and *C* to characterize COVID-19 itself, the contact period *B* to characterize the behavior of the population, the initial latent population *ρ* = *E*_0_/*I*_0_ to characterize undetected community spreading at the onset of the outbreak, the affected population *η* = *N*∗/*N* to characterize containment, and the travel coefficient *ϑ* to characterize spreading through passenger air travel.

### 4.1 The latent and infectious periods *A* and *C* characterize the timeline of the disease

Our sensitivity analysis in Figures 4 and 5 shows the impact of the latent and infectious periods *A* and *C*. Both affect the peak of the infectious population both in time and magnitude. The robust data for the infectious and recovered populations of all 30 Chinese provinces in Figure 9 suggest that the latent period lasts for 2.5 days, followed by the infectious period of 17.8 days. A study of 391 confirmed COVID-19 cases with 1,268 close contacts in Shenzhen found a median incubation period of 4.8 days until the onset of symptoms, a mean time to isolation after the onset of symptoms of 2.7 days or 4.6 days with or without active contact tracing, and a median time to recovery of 20.8 days after the onset of symptoms [5]. These values agree with the reported incubation period of 5.1 days found in 181 confirmed COVID-19 cases outside Wuhan [22] and 5.2 days for the first 425 cases in Wuhan [24]. The total duration from exposure to recovery, (*A* + *C*) of our SEIR model, is 20.3 days, 5.3 days shorted than the reported value of 25.6 for the 391 Shenzhen cases [5]. In our model, the reported 4.8 to 5.2 day incubation periods maps onto the latent period *A* of 2.5 days plus 2.3 to 2.7 days within the infectious period *C* during which the individuals are infectious but still asymptomatic. This period is critical since individuals can spread the disease without knowing it. The contact tracing study postulates that the infectious period *C* begins on day 4.8 with the onset of symptoms, 2.3 days later than in our model, and ends on day 7.3 or 9.4 with or without active contract tracing with the beginning of isolation, 13.0 or 10.9 days earlier than in our model. This implies that the infectious period *C* of our SEIR model is 6.6 and 3.9 times larger than the infectious period of the traced and untraced early isolated population in Shenzhen [5]. This comparison suggests that it is critical to understand how the infectious period is reported, either as a disease-specific parameter or as a medically-modulated exposure time.

### 4.2 The contact period *B* and basic reproduction number *R*_0_ characterize social and political behavior

Our sensitivity analysis in Figures 6, 7, and 8 shows the impact of the contact period *B* or, more intuitively, the basic reproduction number *R*_0_. The basic reproduction number significantly affects the peak of the infectious population both in time and magnitude. The early outbreak data for the infectious populations of all 50 United States in Figure 10 suggest that the contact period is for 3.4 days, resulting in a basic reproduction number of 5.3. For the first 425 cases in Wuhan, the basic reproduction number was estimated to 2.2 [24] and for the 391 cases in Shenzhen, it was 2.6 [5]. A review of the reported basic reproduction numbers for COVID-19 found ranges from 1.40 to 6.49 with a mean of 3.28, values that are larger than those reported for the SARS coronavirus [25]. Huge variations of *R*_0_ values are not uncommon [11]; even for simple diseases and for the 391 cases in Shenzhen, it was 2.6 [5]. A review of the reported basic reproduction numbers for COVID-19 found ranges from 1.40 to 6.49 with a mean of 3.28, values that are larger than those reported for the SARS coronavirus [25]. Huge variations of *R*_0_ values are not uncommon [11]; even for simple diseases like the measles, reported *R*_0_ values vary between 3.7 and 203.3 [10]. Community mitigation and political action can modulate the basic reproduction number *R*_0_ by a variety of measures including active contact tracing, isolation of infectious individuals, quarantine of close contacts, travel restrictions, physical distancing, or total lock down [13]. Importantly, many of the reported values already include the effect of isolation [24] and active contact tracing and quarantine [5]. If we correct our identified basic reproduction number for China in Figure 9 and Table 1 by reducing our identified infectious period of 17.8 days to the time prior to isolation using the correction factors of 6.6 and 3.9 with and without contact tracing, our *R*_0_ values for China would be 1.91 and 3.23 and fall well within the reported range [25]. Our *R*_0_ value for the United States of 5.30 agrees well with the rang of values reported for mathematical model ranging from 1.50 to 6.49 with a mean of 4.20 [25]. Understanding the natural value of *R*_0_–without any mitigation strategy–is critical to predict the endemic equilibrium, interpret herd immunity, and the estimate the fraction of the population that requires vaccination [18].

### 4.3 What’s next?

Current mitigation strategies have the goal to “flatten the curve”, which translates into reducing the number of new infections. As we can see in Figures 6, 7, and 8, we can achieve this goal by reducing the basic reproduction number *R*_0_ = *C*/*B*, which is a direct signature of effective containment measures and drastic behavioral changes that affect a substantial fraction of the susceptible population [13]. By isolating infectious individuals, active contact tracing, and quarantining close contacts, we can reduce the effective infectious period *C*; and by implementing travel restrictions, mandating physical distancing, or enforcing total lock down, we can increase the contact period B [27]. Figure 9 demonstrates that combinations of these measures have successfully flattened the curves in the 30 provinces of China [24]. But the million-dollar questions remains: What’s next? In the very near future, our model has the potential to predict the timeline of the outbreak, specifically, the timing and peak of the infectious population in individual states and countries. This will help us optimize planning and distribute medical resources where needed [17]. In the short term, we could enhance our model to study the effect of different subgroups of the population [5]. This could provide scientific guidelines to gradually relax political measures, for example by releasing different subgroups of the population before others. In the long term, we will need accurate values of the basic reproduction number to estimate the effect of vaccination. This will be critical to design rigorous vaccination programs and prioritize which subgroups of the population to vaccinate first [18]. Naturally, as more data become available, we can train our models more reliably and make more accurate predictions.

### 4.4 Limitations

This study proposes a new strategy to characterize the timeline of COVID-19. While this allows us to estimate the peaks of the outbreak in space and time, we need to be aware that this study uses a simple model to characterize a complex infectious disease about which we still know very little to this day. Importantly, we have to be cautious not to overstate the results. Specifically, our study has several limitations: First, our mathematical model does not account for asymptomatic cases. Little is known about the fraction of asymptomatic or mildly symptomatic individuals but early studies suggest that up to 25% of individuals have gone from susceptible to recovered without having ever been reported as infectious. Second, the classical SEIR model does not distinguish between asymptomatic infectious in the first days of the disease and symptomatic infectious in the later days. Knowing more about this group and modeling appropriately is critical to accurately estimate the impact of community spreading and mitigation strategies to reduce it. Third, while the initial infectious group *I*_0_ can be reasonably well approximated from the reported active cases and the initial recovered group *R*_0_ is likely zero, the initial exposed group E0 is really unknown and can hugely effect the outbreak dynamics as the sensitivity study in Figure 3 and the data for China and the United States in Figures 9 and 10 show. We decided to include this effect through the initial latent population *ρ* to highlight this effect, but more data are needed to better estimate the size of this group. Fourth and probably most importantly, the major variable we can influence through social and political measures is the basic reproduction number *R*_0_, or rather the interplay of the contact period *B* and infectious period *C*. Obviously, we do not know the true *R*_0_, nor can we measure it at this stage of the outbreak, where every state, province, or country has implemented different measures to modulate the local outbreak dynamics. Nonetheless, our study shows that estimating *R*_0_ is important to quantify if and how different political counter measures work and to predict the timeline of the infectious population under no, moderate, and massive political action. Finally, our network model only provides rough mobility estimates from air travel statistics. To more accurately simulate the spreading of COVID-19, we could gradually refine our network and include more granular mobility patterns, for example from cell phone data.

## 5 Conclusion

The precise timeline of COVID-19, its basic reproduction number, and the effect of different mitigation strategies remain incompletely understood. Here we combined data from the outbreak in China with data from the early stages of the outbreak in the United States to identify the latent, contact, and infectious periods and the basic reproduction number of COVID19. To quantify the outbreak dynamics, we integrated a global network model with a local epidemic SEIR model and solved the resulting set of coupled nonlinear equations using a Newton-Raphson scheme. For the outbreak in China, in n = 30 provinces, we found a latent period of 2.6 days, a contact period of 1.5 days, and an infectious period of 17.8 days. For the early stages of the outbreak in the United States, in n = 50 states, we found a contact period of 3.4 days and a travel coefficient of 0.42. Our network model predicts that–without the massive political mitigation strategies that are in place today–the United states would have faced a basic reproduction number of 5.3±0.95 and a nationwide peak of the outbreak on May 10, 2020 with 3 million infections. Our results suggest that mathematical modeling can help estimate outbreak dynamics and provide decision guidelines for successful outbreak control. Our model has the potential to quantify the impact of community measures and predict the effect of relaxing total lock down, shelter in place, and travel restrictions for low-risk subgroups of the population or for the population as a whole.

## Data Availability

all data are publicly available as cited in the references

## Acknowledgements

This work was supported by a Stanford Bio-X IIP seed grant to Mathias Peirlinck and Ellen Kuhl and by a DAAD Fellowship to Kevin Linka.

